# The effect of multigeneration history of suicidality on offspring’s neurodevelopment outcomes: evidence from the Adolescent Brain and Cognitive Development (ABCD) cohort

**DOI:** 10.1101/2022.05.24.22275547

**Authors:** Xue Wen, Diyang Qu, Guowei Wu, Dongyu Liu, Yuanyuan Wang, Zaixu Cui, Xiaoqian Zhang, Runsen Chen

## Abstract

**Background:** Parent-child transmission of suicidal behaviors has been widely elucidated, while the three-generation family suicide risk paradigm remains to be explored. This longitudinal study aimed to examine the influences of family history of suicidality (FHoS) among two prior generations on offspring’s neurodevelopment.

**Methods:** We conducted a retrospective, longitudinal study using the Adolescent Brain Cognitive Development (ABCD) study data collected from 2016 to 2021. Participants were allocated into four groups according to their parents’ (Generation 1 [G1]) and grandparents’ (Generation 2 [G2]) history of suicidality (G1−G2−; G1+G2−; G1−/G2+; G1+/G2+). We estimated adjusted associations between FHoS and offspring’s suicide ideation (SI), psychopathology, impulsivity and brain cortical volumes while controlling for age, sex, parental education, household income and marital status.

**Findings:** A total of 11,875 children aged 9-10 years were observed from baseline to 3-year follow-up. Compared to G1-G2-, higher odds of SI were observed for G1-G2+ (OR=1·99, 95% CI [1·54-2·56]) and G1+G2+ (2·25 [1·46-3·47]) by child-report. Higher odds of SI were also observed for G1+G2- (1·54 [1·12-2·12]), G1-G2+ (2·57 [1·89-3·48]) and G1+G2+ (2·70 [1·60-4·56]) by caregiver-report. Higher odds for psychopathology were also observed (1·47 [1·11-1·96]; 3·33 [2·57-4·33]; 5·44 [3·42-8·66]), while higher family suicide risk was associated with high impulsivity (B=1·32 [0·48-2·17]; 2·24 [1·32-3·15]; 2·26 [0·47-4·05]). Offspring in G1+G2-had higher cortical volumes in 12 brain regions, including the bilateral insula, temporal regions and occipital regions, which were also significantly associated with their lifetime SI.

**Discussion:** A cumulative risk pattern of FHoS in two prior generations was found for offspring’s neurodevelopmental outcomes. Earlier preventive interventions are warranted to weaken the familial transmission of suicidal risk.

## Introduction

Suicide ideation (SI), which is always seen as the precursor of suicide attempt (SA),^1^ has been sharply increasing among children over the past decades.^2^ The global prevalence of SI among adolescents were 10%-17% from 2003 to 2015.^3^ Noteworthy, SI also has a long-lasting effect on children’s neurodevelopmental outcomes, including cognitive and emotional abilities.^4–8^ This may further transmit to adulthood and even lead to a high mortality risk,^9^ and a staggering toll on global public health. A 50-year meta-analysis systematically reviewed the multi-dimensional risk factors of SI,^10^ which highlighted the importance of family history of suicidality (FHoS). Additionally, child with a FHoS were more vulnerable compared to adolescents and adults^11,12^.

Several frameworks was been used to explain this transmission, including shared genetic predispositions,^13^ contagions,^14^ adverse environmental circumstances and imitation.^15^ Abundant population-based studies have provided detailed evidence for these explanations.^16–26^ Additionally, a neuroimaging study further explored the specific phenotypes resulted by FHoS, including reduced volumes in bilateral temporal regions, right dorsolateral prefrontal cortex and left putamen.^27^ However, the majority of previous intergenerational studies have focused on the parent-to-child transmission of suicidality, while the impact of other family members were commonly neglected. Children’s development is deeply rooted in the whole family and is also closely linked to other family members, especially for grandparents, who play a crucial role in children’s development both directly and indirectly.^28^ For instance, grandparents’ mental disorders could act on their grandchildren independently,^29^ or through the intermediate influence of parents.^30^ Nonetheless, to what extent the FHoS within three generations influences offspring’s SI and other neurodevelopment outcomes remains strikingly unknown, which greatly hinders the design of more cost-effective interventions in early stage.

Therefore, this study aimed to explore the longitudinal effects of FHoS within two prior generations on children’s self-report SI, their caregiver’s report of SI, and other development outcomes, including psychopathology and impulsivity. While the potential protective roles of sex and SES among these associations were also explored. We further investigate whether offspring’s cortical volumes were associated with grandparental suicidality, and its correlations to their lifetime SI.

## Methods

### Data sources

We conducted a retrospective, population-based, longitudinal study by leveraging large sample data in the Adolescent Brain and Cognitive Development (ABCD) study.^31^ From the National Institutes of Health (NIH) website, we obtained data of “Curated Annual Released 4.0”, which included 11,875 children recruited from 22 sites across the United States in the baseline (2016-2018, aged 9-10 years), 11,224 individuals in the 1-year follow-up (2017-2019, aged 10-11 years), 10,413 individuals in the 2-year follow-up (2018-2020, aged 11-12 years) and 6,250 individuals in the 3-year follow-up (2019-2021, aged 12-13 years). The present study focused on the data at all these four time points. All written informed content from parents and verbal assent from children were available in this analysis. All procedures were approved by a centralized institutional review board (IRB) from the University of California, San Diego IRB. Each study site obtained approval from their local IRBs. During data collection and analysis, all the ethical regulations have been complied with.

### Study sample

We included data of 11,875 children from baseline to 3-year follow up. Herein, demographic information was obtained from the caregiver-report demographics battery, including age, sex at birth, race/ethnicity, household marital status, parental education level and combined household income. The highest value of parental education was selected, then the original 21 different response options were re-coded into 5 levels (Less than High School, High School Graduate/GED, Some College, Associate’s/Bachelor’s Degree and Postgraduate Degree). It was further re-coded into 2 categories (Less than college, College and above) for preliminary analysis. The original nine different response options of household income were also re-coded into three levels (<50K, ≥50K&<100K, ≥100K) for further analysis. These transformations were aimed to simplify our statistical models without losing nuance and have already been implemented in previous studies^32,33^. Since parental education level and household income are widely used indicators of family socioeconomic status (SES)^34^, they were combined to define low SES and high SES (appendix).

### Exposures

The Family History Assessment Module Screener (FHMH-S) questionnaire reports the presence/absence(+/-) of a range of mental health symptoms including SA or suicide death in all 1st and 2^nd^ degree “blood relatives” of the child^35^. The total score of FHoS was obtained by summing the histories from paternal grandparents, maternal grandparents and parents. The types of combination were further considered to generate a new 2×2 variable (G1-G2-; G1+G2-; G1-G2+; G1+G2+), which reflects both the density and distance of offspring suicide exposure genetically and environmentally. Among these variables, G1 represents the presence of grandparents’ FHoS, and G2 represents of the presence of parents’ FHoS (appendix p 1).

### Outcomes

The suicide module of the computerized Kiddle Schedule for Affective Disorders and Schizophrenia (KSADS, Lifetime version) was used to assessed participants’ SI at present or in the past. Specifically, SI includes passive SI, non-specific active SI, active SI with method, active SI with intent and active SI with plan. In addition, the youth-report and caregiver-report version of KSADS were used simultaneously. The youth-report data was collected at all four data collection points, while the caregiver-report data was only available in baseline and 2-year follow-up assessment.

Offspring problematic behaviors over the past 6 months were accessed by parent-report data from the Child Behavior Checklist (CBCL; age 6 to 18 form).^36^ The CBCL is a 3-point Likert-scale with response options of 0 (“not true”), 1 (“somewhat or sometimes true”), and 2 (“very true or often true”). It is used to judge the extent to which the problematic behaviors characterize their children. Then the total score was re-coded as being within (<60) versus above the threshold for borderline clinical significance, as previously defined^37,38^. Data from baseline, 1-year, 2-year and 3-year follow-up assessment was selected for further analysis.

Offspring trait impulsivity was measured by the short version of the UPPS-P self-report scale.^39^ Children rated on 4-point Likert-type scale among 1 (“Not at all like me”), 2 (“Not like me”), 3 (“Somewhat like me”), or 4 (“Very much like me”). In addition, the total score was converted to z-scores using a Fisher’s R to Z transformation. Children with a total score lower or higher than 3 SDs from the total group mean were excluded as outliers. Data from baseline and 2-year follow-up assessment was selected for further analysis. As for offspring brain structures, only baseline assessment was selected.

### Statistical Analysis

All statistical analyses were performed using SPSS (version 26.0). In the preliminary analyses, the prevalence of offspring SI over the 4 years among different demographic backgrounds and family suicidal backgrounds was first examined using cross-tabulations. We then estimated the predictive associations between the factors above and offspring SI in a series of binomial regression models. Odds ratios (ORs) and confidence intervals (CIs) were presented to explain the results, especially for the association between number of FHoS and offspring SI reported by child and caregiver separately. We then conducted a series of analyses around different types of FHoS (G1-G2-; G1+G2-; G1-G2+; G1+G2+). Propensity-score-matched (PSM) analysis was used to account for the sample size differences in the G1-G2- and other three groups and to minimize selection bias. Then generalized estimating equations (GEE) were used to obtain the adjusted relative risks of SI, psychopathology and impulsivity in preadolescents with FHoS. Multi-variable regression models were constructed a priori to adjust for the effect of important confound variables: age, sex at birth, race/ethnicity, household marital status, parental education level, household income and site. We continued to test whether the child’s sex at birth and SES modified the associations between FHoS and child prevalence of SI and other development outcomes in the final interaction analysis. The detailed descriptions of the methods above were shown in the appendix. Subgroup analysis were further conducted between G1+G2- and G1-G2-. FreeSurfer v5.3 (http://surfer.nmr.mgh.harvard.edu) was used to process the locally acquired T1-weighted images, and estimate global and regional gray matter volume, while Desikan-Killiany atlas was used to generate 74 cortical parcellations within each hemisphere.^40^ The volumes in left and right hemispheres were added for each region. In the general linear models, which included covariates selected a priori, t-statistics was estimated across these regions. Total volumes of the interested regions were final calculated, then correlated to offspring SI among G1+G2-.

## Results

The total study sample of 11,875 children were observed over 4 years. Altogether, 2,309 (19.4%) children self-reported the presence of SI. Herein, female (OR=1.13, 95% CI [1.03-1.24]), unmarried families (OR=1.18, 95% CI [1.05-1.33]), low household income (OR=1.20, 95% CI [1.04-1.40]) and high parental education level (OR=1.16, 95%CI [1.01-1.32]) were associated with significantly increased odds of offspring SI. Specifically, a dose-response relationship was observed between number of FHoS and offspring SI, as the number of FHoS increase, the relative odds of offspring SI also increase. A similar, although less consistent pattern is shown in SI reported by caregiver, with only 1,420 (12.0%) children were reported to have SI (Table 1).

**Table 1:** Characteristics of offspring with SI and logistic regression outcomes.

| Characteristic | Child-report(N=2309) |  |  | Caregiver-report(N=1420) |  |  |
| --- | --- | --- | --- | --- | --- | --- |
|  | SI<sup>a</sup> | OR(95%CI) | p value | SI<sup>b</sup> | OR(95%CI) | p value |
| Sex at birth |  |  |  |  |  |  |
| Male | 1150(49.8%) | 1(ref) |  | 839(59.1%) | 1(ref) |  |
| Female | 1158(50.2%) | 1.13(1.02-1.25) | 0.017 | 581(40.9%) | 0.74(0.65-0.84) | <0.001 |
| Household married |  |  |  |  |  |  |
| Married | 1463(63.4%) | 1(ref) |  | 913(64.3%) | 1(ref) |  |
| Unmarried | 831(36.0%) | 1.19(1.02-1.39) | 0.03 | 500(35.2%) | 1.21(1.04-1.42) | 0.016 |
| Household income |  |  |  |  |  |  |
| ≥100k | 811(38.3%) | 1(ref) |  | 521(39.8%) | 1(ref) |  |
| ≥50k<100k | 613(29.0%) | 1.12(0.99-1.27) | 0.085 | 399(30.5%) | 1.03(0.85-1.25) | 0.786 |
| <50k | 693(32.7%) | 1.21(1.06-1.37) | 0.004 | 389(29.7%) | 1.11(0.95-1.29) | 0.190 |
| Parental highest education |  |  |  |  |  |  |
| Less than college | 626(27.2%) | 1(ref) |  | 339(23.9%) | 1(ref) |  |
| College and above | 1679(72.8%) | 1.18(1.02-1.35) | 0.023 | 1079(76.1%) | 1.35(1.13-1.61) | 0.001 |
| Number of FHoS |  |  |  |  |  |  |
| N=0 | 1836(87.8%) | 1(ref) |  | 1034(83.3%) | 1(ref) |  |
| N=1 | 210(10.0%) | 1.42(1.20-1.68) | <0.001 | 171(13.8%) | 2.12(1.76-2.56) | <0.001 |
| N≥2 | 44(2.9%) | 2.49(1.69-3.69) | <0.001 | 37(3.0%) | 3.89(2.58-5.88) | <0.001 |

Ref=reference. <sup>a</sup>Total number of offspring with SI was obtained from baseline to three-year follow up by child-report.
<sup>b</sup>Total number of offspring with SI was obtained from baseline and two-year follow up by caregiver-report.

Baseline characteristics of the population after PSM analysis were summarized in Table 2. The present study included 1,848 out of 11,875 children for further GEE analysis, in which the reference group (G1-G2-, N=924) were 1:1 matched with the other 3 group. Among them, 887 (48.0%) were female, and the mean (SD) age was 9.9 (0.6) year. Other demographic characteristics were distributed across the 4-risk types.

**Table 2:**
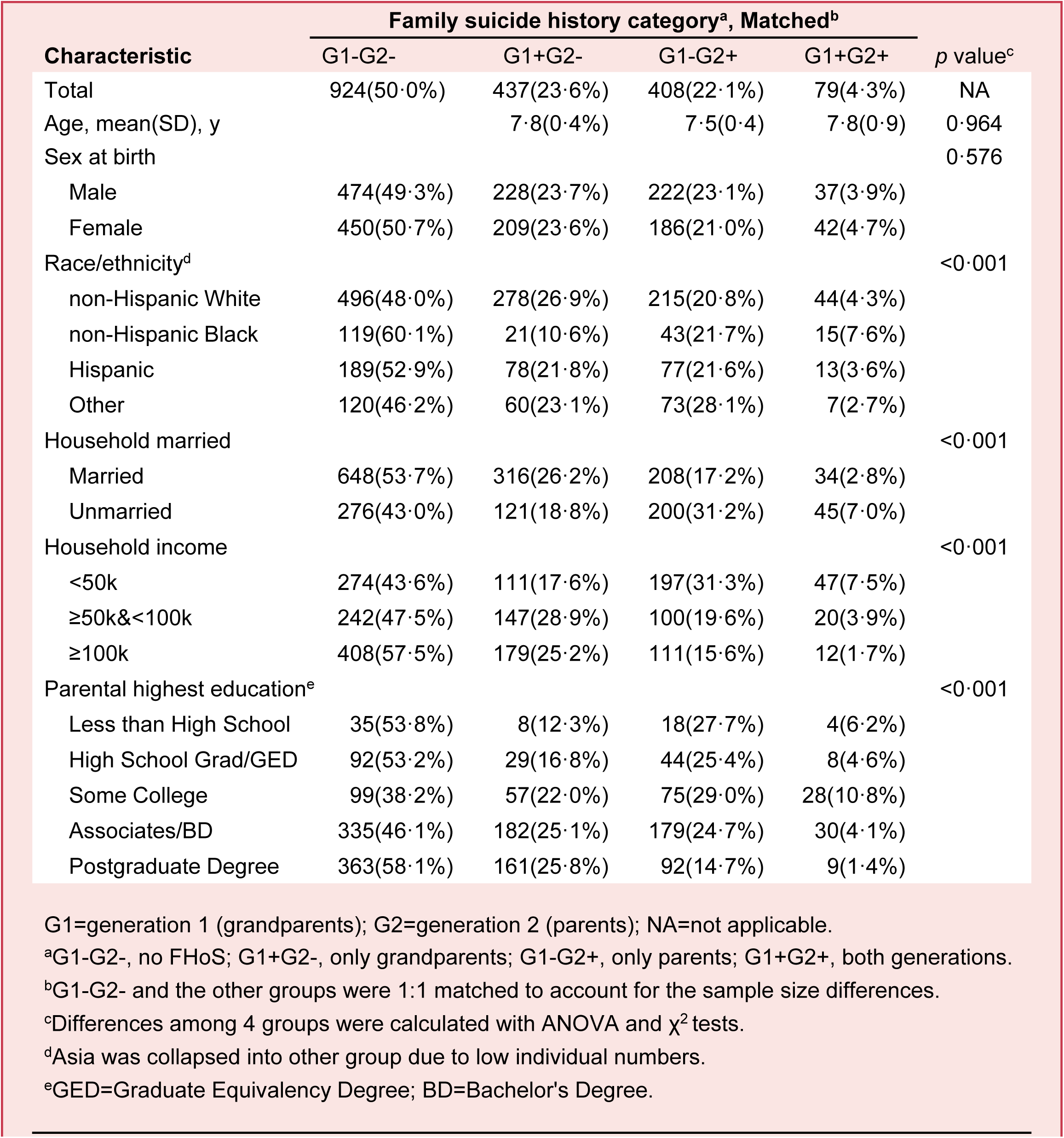
Demographic characteristics of sample by 4-risk types of FHoS after PSM.

GEE models revealed that parental and grandparental SA or suicide death is significantly associated with increased odds of subsequent SI in offspring. By child reports, the odds of SI were 1.99 (95%CI [1.54-2.56]) times higher in G1-G2+ and 2.25 (95%CI [1.46-3.47]) times higher in G1+G2+ compared to G1-G2-after adjusted. By caregiver reports, the odds of SI were 1.54 (95%CI [1.12-2.12]) times higher in G1+G2-, 2.57 (95%CI [1.89-3.48]) times higher in G1-G2+ and 2.70(95%CI [1.60-4.56]) times higher in G1+G2+ after adjustment. Furthermore, the prevalence of offspring SI reported by themselves decreased among the 4 years but with some dampening of the trend (Fig 1). For example, the prevalence of SI was 10.8% in baseline, 8.8% in 1-year follow up, 8.5% in 2-year follow up while 14.8% in 3-year follow up. Moreover, the main effect of sex and SES is not significant, yet sex at birth (χ^2^=34.27; p<0.001) and SES (χ^2^=37.30; p<0.001) moderated the association between FHoS and offspring SI by child reports. While by caregiver reports, boys (mean, 0.15; SE, 0.014) had higher prevalence of SI than girls (mean, 0.11; SE, 0.011). Sex at birth (χ^2^=50.16; p<0.001) and SES (χ^2^=37.65; p<0.001) also moderated the association between FHoS and offspring SI (Fig 2).

**Figure1:**
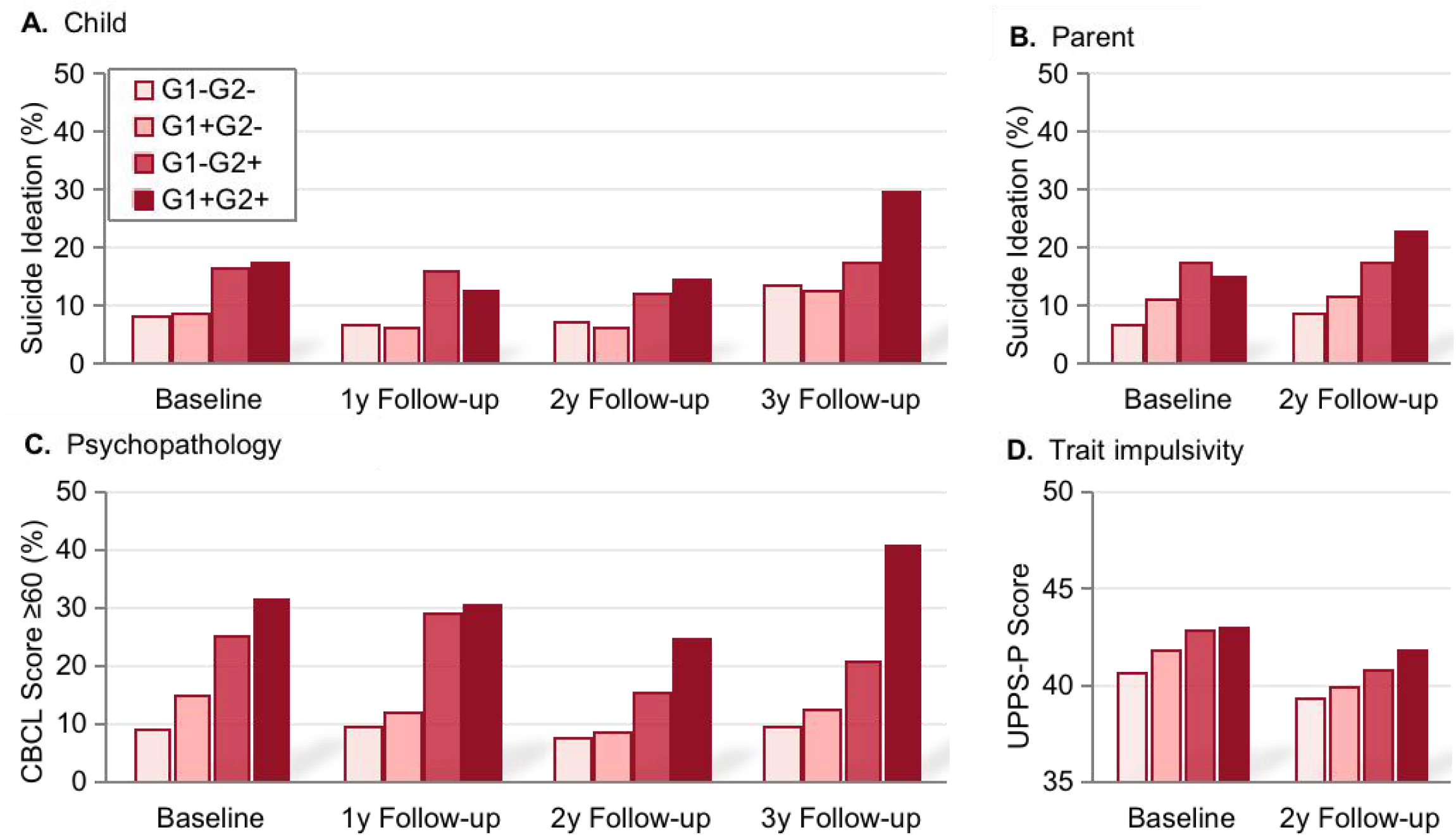
Offspring developmental outcomes among 4 years by family suicide risk

**Figure2:**
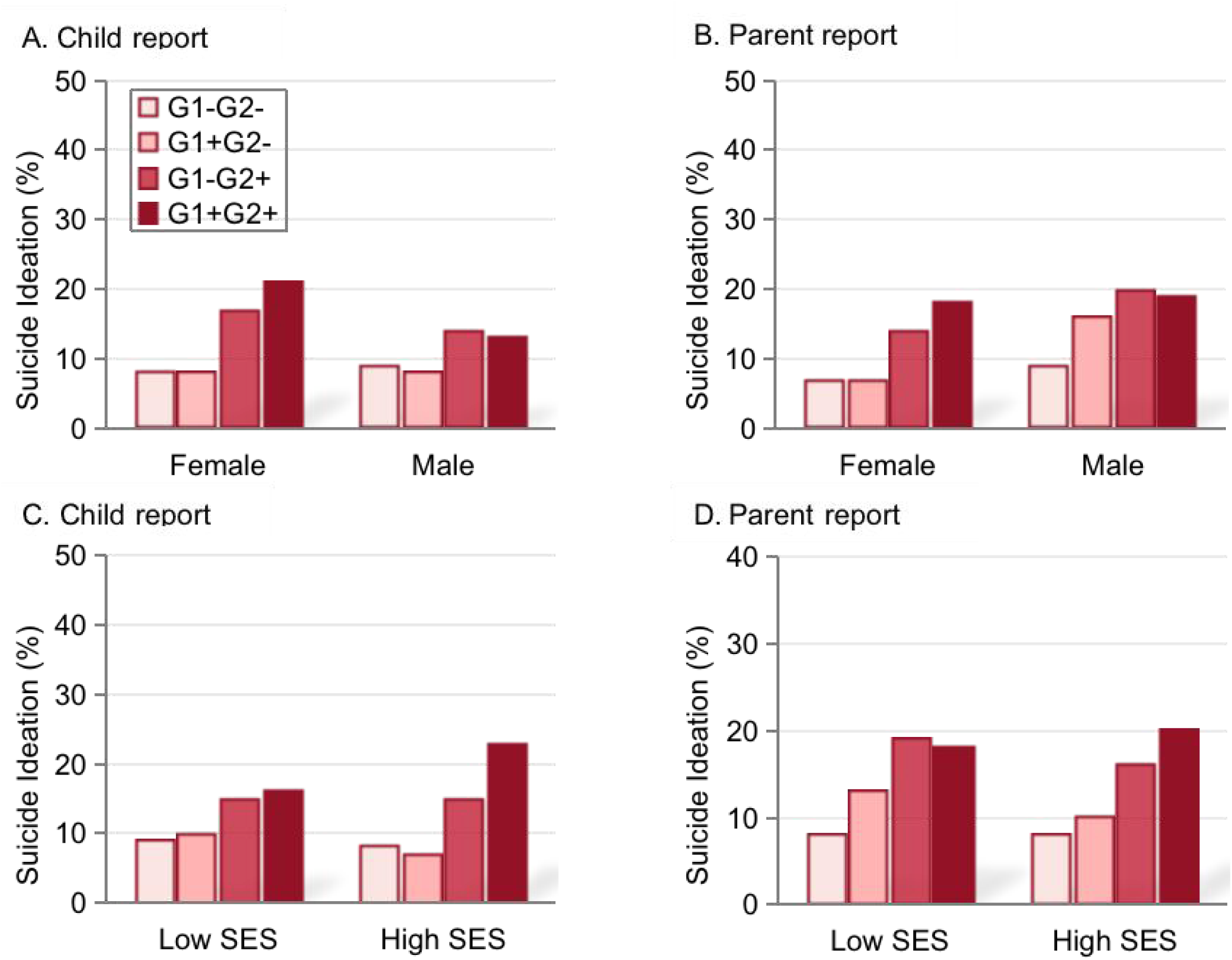
Sex and SES moderated the association between FHoS and offspring’s SI

FHoS also predicted clinically relevant psychopathology in offspring (Table 3). Higher likelihood to have psychopathological behaviors was observed with increased levels of FHoS. The odds ratio is 1.47 (95%CI [1.11-1.96]) in G1+G2-, 3.33(95%CI [2.57-4.33]) in G1-G2+ and 5.44 (95%CI [3.42-8.66]) in G1+G2+ compared to G1-G2-. There was no time effect in the analysis (Fig 2). Additionally, girls (mean, 50.07; SE, 0.55) performed more psychopathic behaviors than boys (mean, 47.18; SE, 0.52), while offspring in low SES (mean, 49.94; SE, 0.49) also showed more psychopathic behaviors than in high SES (mean, 47.19; SE, 0.62). Sex at birth (χ^2^=142.52; p<0.001) and SES (χ^2^=142.52; p<0.001) moderated the association between FHoS and offspring psychopathology (Fig 3).

**Table 3:**
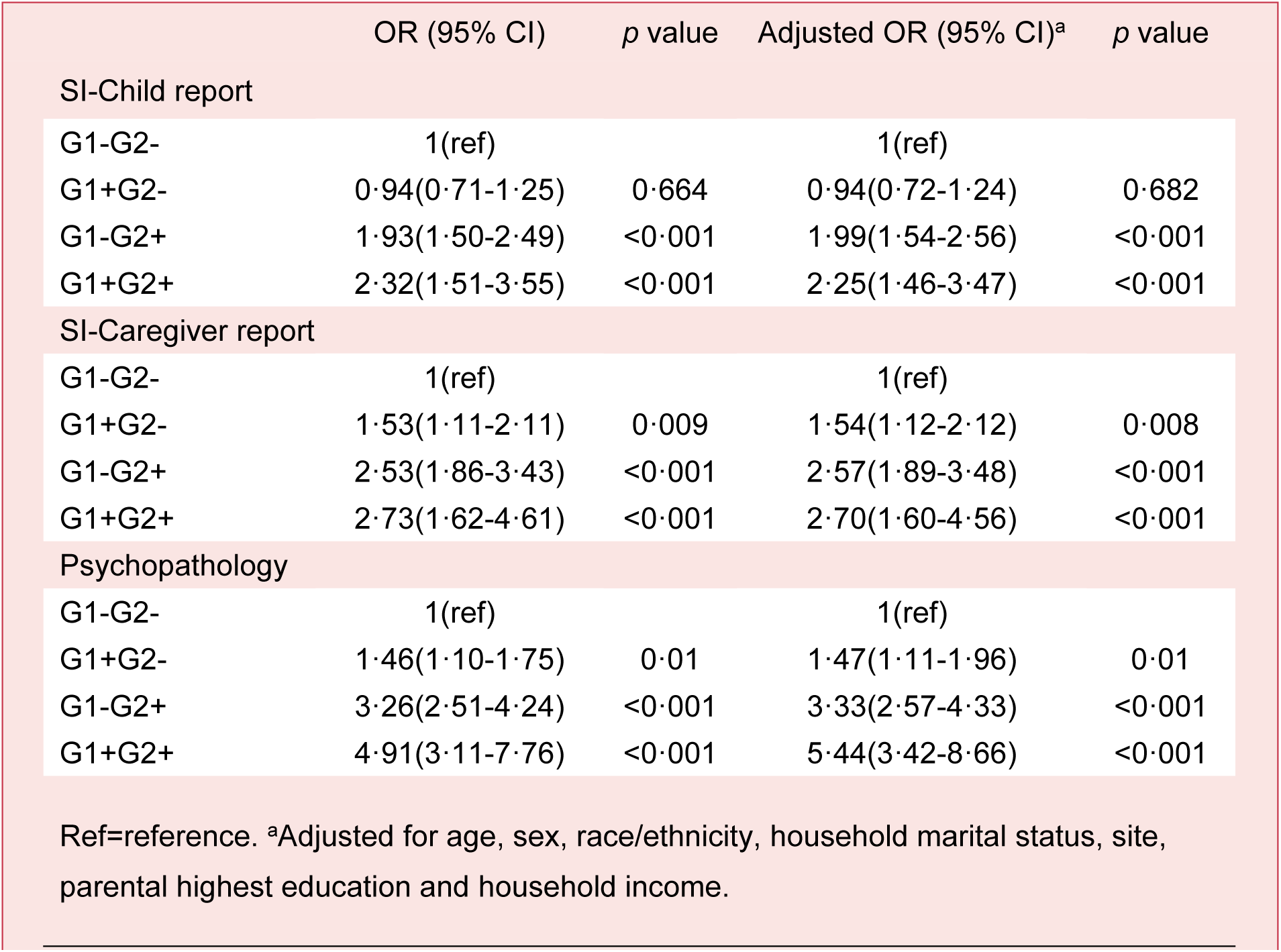
Increased family suicide risk on offspring’s SI and psychopathology.

**Figure3:**
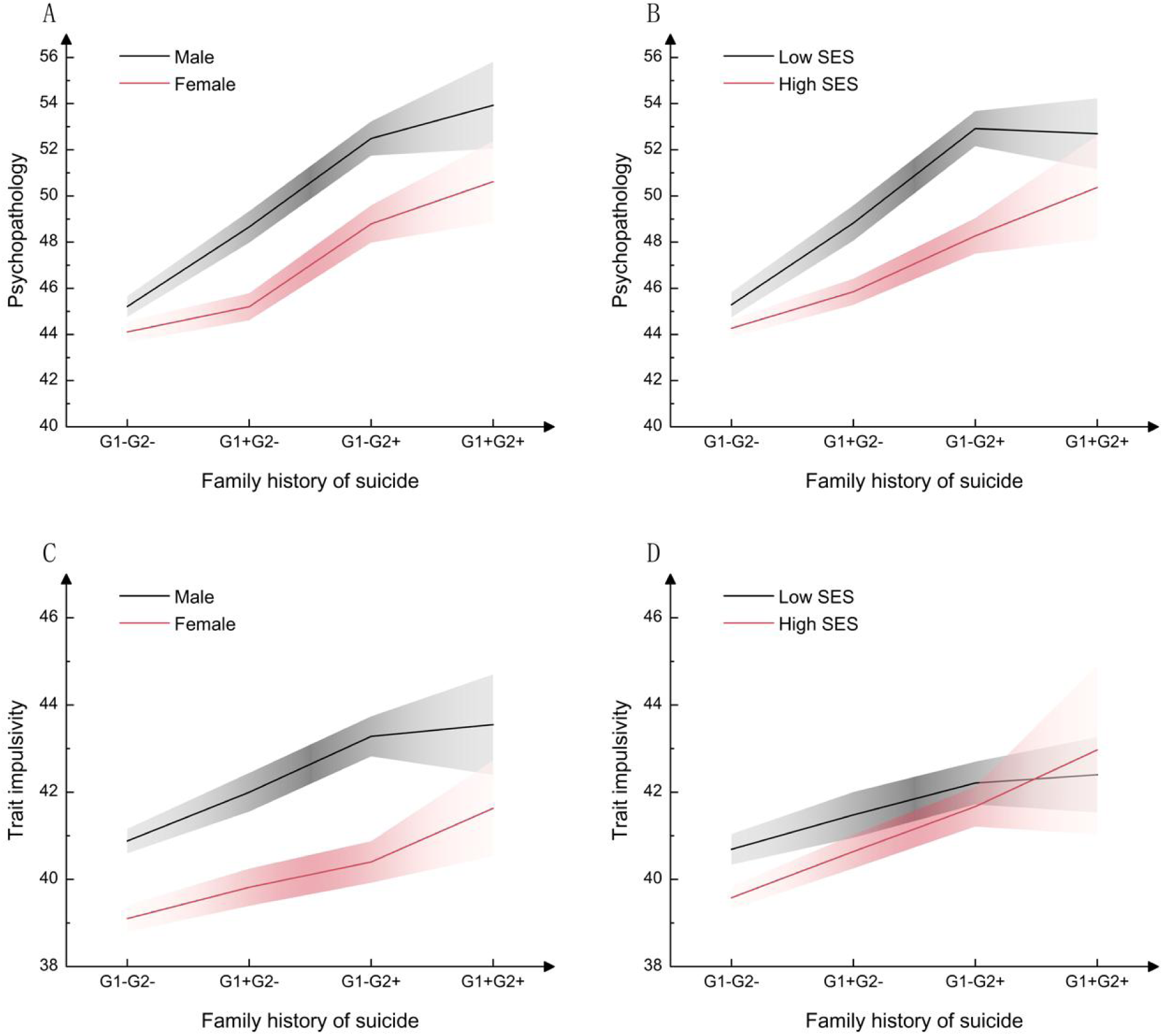
Sex and SES moderated the association between FHoS and offspring’s psychopathology and impulsivity

There was also evidence for higher impulsivity in children with FHoS (Table 4). Compared to G1-G2-, G1+G2- (B=1·32, 95%CI [0·48-2·17]), G1-G2+ (B=2·24, 95%CI [1·32-3·15]) and G1+G2+ (B=2·26, 95%CI [0·47-4·05]) were associated with higher impulsivity. Meanwhile, time effect was observed since lower impulsivity is measured in 2-year follow up (mean, 40.49; SE, 0.29) than baseline (mean, 42.05; SE, 0.27). There is no FHoS by time interaction in the model. Furthermore, boys (mean, 42.43; SE, 0.34) had higher trait impulsivity than girls (mean, 40.24; SE, 0.33), despite that the main effect of SES is not significant. Sex at birth (χ^2^=79.15; p<0.001) and SES (χ^2^=39.98; p<0.001) moderated the association between FHoS and offspring trait impulsivity (Fig 3).

**Table 4:** Increased family suicide risk on offspring’s impulsivity.

|  | B (95% CI) | *p* value | Adjusted B(95% CI)<sup>a</sup> | *p* value |
| --- | --- | --- | --- | --- |
| Trait impulsivity |  |  |  |  |
| G1-G2- | 0(ref) |  | 0(ref) |  |
| G1+G2- | 1.30(0.44-2.16) | 0.003 | 1.32(0.48-2.17) | 0.002 |
| G1-G2+ | 2.20(1.28-3.12) | <0.001 | 2.24(1.32-3.15) | <0.001 |
| G1+G2+ | 2.38(0.55-4.21) | 0.011 | 2.26(0.47-4.05) | 0.014 |
| Ref=reference. <sup>a</sup>Adjusted for age, sex, race/ethnicity, household marital status, site, parental highest education and household income. |  |  |  |  |
| **Table 4: Increased family suicide risk on offspring's impulsivity** |  |  |  |  |

In comparison to G1-G2-, 12 significant differences across the 74 cortical regions were found after false discovery rate (FDR) correction, all indicating increased brain volumes among children in G1+G2-, of which the strongest difference was the volume of the superior segment of the circular sulcus of the insula (*t*=12.89, P_FDR_<0.001). Meanwhile, total volumes of these 12 regions were negatively correlated to offspring SI both by child-report (r=-0.136, P=0.005) and by caregiver-report (r=-0.115, p=0.018) (Fig 4).

**Figure4:**
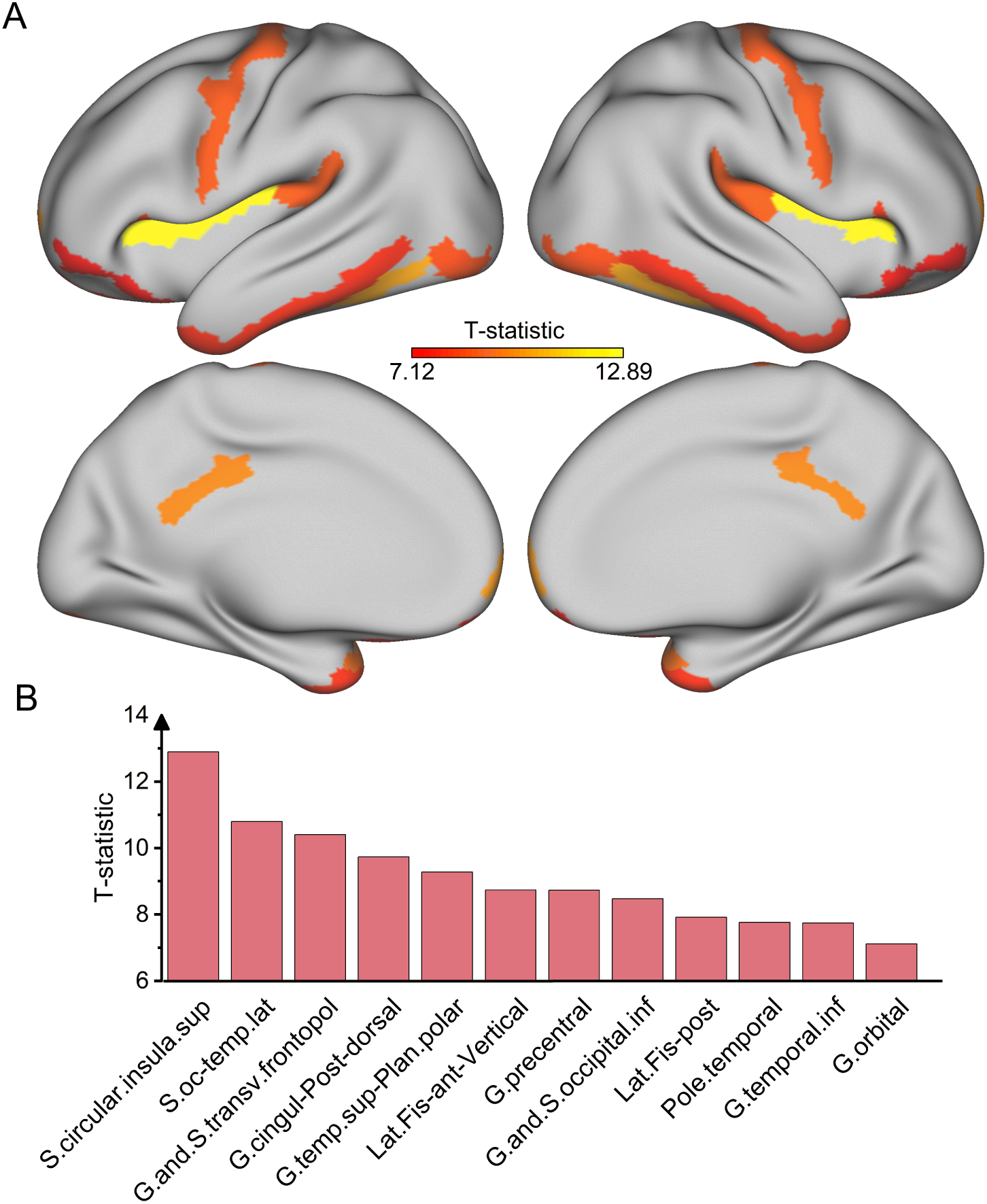
Representative regions with higher volumes in offspring with grandparental suicidality. **A**. Increased cortical volumes were found among G1+G2 compared to G1-G2- in 12 regions (sum of left and right hemisphere), while covarying for age, sex, race/ethnicity, household marital status, parental highest education and household income. These representative regions have been projected onto the surface of the brain for better visualization. **B**. Representative regions and their corresponding T values are shown in the bar chart. S.circular.insula.sup=superior segment of the circular sulcus of the insula; S.oc-temp.lat= lateral occipito-temporal sulcus; G.and.S.transv.frontopol=transverse frontopolar gyri and sulci; G.cingul-Post-dorsal=posterior-dorsal part of the cingulate gyrus; G.temp.sup-Plan.polar=planum polare of the superior temporal gyrus; Lat.Fis-ant-Vertical= vertical ramus of the anterior segment of the lateral sulcus; G.precentral=precentral gyrus; G.and.S.occipital.inf=inferior occipital gyrus and sulcus; Lat.Fis-post= posterior ramus of the lateral sulcus; G.temporal.inf=inferior temporal gyrus; Pole.temporal=temporal pole; G.orbital= orbital gyri.

## Discussion

This study addresses the gap of multigeneration transmission of suicidal risks through three key findings. Firstly, the increasing number of family history of suicidality across generations predicts higher level of offspring’s SI and development outcomes. Specifically, having at least one grandparent and parent with SA or suicide death indicates the highest prevalence of SI and the highest level of psychopathology and impulsivity in offspring. Secondly, sex at birth and SES moderated the relationship between FHoS and offspring’s SI, problematic behaviors and impulsivity. Thirdly, higher regional cortical volumes in offspring with grandparental suicidality were found, especially for insula, while the sum of these 12 regions were negatively associated with offspring SI. Together, these results provide evidence for the effect of FHoS on offspring’s neurodevelopment, as well as the brain-structure-related determinants of the intergenerational transmission of suicidality.

Our findings of the highest risk for offspring’s SI, psychopathology and impulsivity in G1+G2+ clarify how the interaction between parental and grandparental SA or suicide death influences offspring mental health status. Since the addictive model and the multiplicative model are widely used to assess complex interactions of two (or more) different exposures,^41^ while there is also evidence suggesting that risk patterns for multigenerational psychiatric disorders follow the addictive model.^29,42–45^ Thus, our study validates the cumulative risk pattern for FHoS. On the basis of parent-child transmission of suicidality, we found that exposure to parental suicidality was not the most dangerous risk factor. Indeed, the longer the time distances spanned through multi-generations, the more the child is influenced by FHoS, as the highest prevalence of SI was reported among preadolescents in G1+G2+, which still increased continuously during the transition to adolescents.

The intergenerational transmission could be explained by shared genes, and a recent molecular genetic study have already identified single nucleotide polymorphisms for the transmission of SA.^46^ Therefore it is reasonable to infer that having multiple generations attempted or suicide death may prompt higher genetic loadings for suicide. Despite the genetic predispositions, the transmission of suicide could also be linked with the transmission of psychopathology and impulsivity,^47–49^ which are always seen as predictors of SI in children and adolescents.^50–53^ Since children with FHoS may have an earlier understanding of suicide and related concepts, and develop the impulsive personality traits and psychopathological behavior characteristics when spending time with their grandparents and parents. Otherwise, offspring’s trait impulsivity tend to decrease over the course of their development, as shown in a prior study.^54^

Our findings validate sex-specific and SES-specific influence in the pathway linking FHoS to offspring’s mental health status. By child reports, our work demonstrated that females were at a particularly higher risk for having SI when exposed to FHoS, which mirrors a previous study.^17^ This is partly because females were more affected by maternal mental health problems since they have more emotional interactions with their mothers.^55^ On the contrary, males were more likely to exhibit psychopathological behaviors and trait impulsivity when exposed to FHoS in our analysis, which could be explained by stronger heritability of problematic performances,^56^ and impulsivity in males.^57^ Meanwhile, offspring with a low SES tends to be more vulnerable to FHoS. Since individuals with a lower SES tend to receive less developmental resources and experience more negative conditions for maintaining mental health,^58,59^ they are more likely to develop mental health problems and psychopathological behaviors.^60,61^ There are also strong intergeneration associations in SES which might also explain our results.^62^ Future research should further consider the potential impact of different geographic, ecological and cultural contexts, especially for the cultural differences between the West and the East.^63^

Our findings demonstrate the role of the insula and other cortical regions in the intergenerational transmission of suicidality. Prior studies have elucidated neuroanatomic regions involved in suicidality, while the insula had been identified as a key neurobiology basis of suicide^64–69^. As previously suggested, the insula was involved in emotion and interoceptive processing,^70^ cognitive control and self-recognition^71^, thus leading to a crucial role in SI^69^. We inferred that the effect of grandparental suicidality on offspring SI might be associated with the change of insula. While future studies need to examine our findings through a longitudinal design in order to access causal effects and identify reliable biological risk markers for suicidality.

Nevertheless, this study also has several limitations. Firstly, due to our novel grouping design for FHoS, the number of SA in each group was too small to be included, while the incidence of G1+G2+ was also relatively low. Therefore, future studies should validate our findings within a large cohort. Secondly, PSM was used to make the statistical analysis of the differences between groups more reasonable, but it may also make the G1-G2-group deviate from its true distribution. Moreover, the results of this study showed partly separate trends under parent- and child-reported SI, consistent with prior studies. Future studies could develop specific algorithms to integrate the results of both interviews to obtain a diagnosis closer to clinicians.^72^ Finally, other protective factors between FHoS and offspring psychopathological symptoms and disorders should be explored in future. Notwithstanding these drawbacks, our study has the following clinical implications. Herein, early identification of FHoS as suicide risk factor among preadolescent is imperative to break the cycle of suicide transmission, which is helpful to the design of cost-effective interventions in early stage, though there were striking few interventional studies related.^73^ Future interventions should also consider children’s higher psychopathology and impulsivity at the same time, and the way to reduce these symptoms. Relatively, psychopathological problems in offspring with a FHoS could also serve to inform interventions for these high-risk families.^74^ Otherwise, inclusion of grandparental suicidal experiences also helped detecting candidates for biomarkers studies earlier.^45^

To conclude, the results of the study show transmission of suicidality in three-generation with an addictive pattern, indicating that the higher psychopathology and impulsivity should be alerted imperatively in clinical settings due to their predictive effect on offspring SI with FHoS. Our study also supports that family history approach is efficient in mapping intergeneration family risk to a population-based study, which is generalizable across different sociodemographic characteristics.^42^ Thus future neuroimaging studies could be conducted to detect biomarkers for FHoS at an early stage.

## Data Availability

All data produced are available online at http://dx.doi.org/10.15154/1523041.

## Contributors

WX, QD, CZ, ZX and RC designed the study. WX and WG assisted data cleaning. WX, QD analysis the data. WX, DQ, CZ, RC, ZX, WY interpret data. WX and QD wrote the first draft of the manuscript. WX and WG made the figures. RC, LD, CZ, QD revised the manuscript critically. All authors contributed feedback and approved the final manuscript.

## Declaration of interests

We declare no competing interests

## Acknowledgments

Data used in the preparation of this article were obtained from the ABCD Study (https://abcdstudy.org), and are held in the NIMH Data Archive (NDA). This is a multisite, longitudinal study designed to recruit more than 10,000 children age 9-10 and follow them over 10 years into early adulthood. The ABCD Study is supported by the National Institutes of Health (NIH)and additional federal partners under award numbers U01DA0401048, U01DA050989, U01DA051016, U01DA041022, U01DA051018, U01DA051037, U01DA050987, U01DA041174, U01DA041106, U01DA041117, U01DA041028, U01DA041134, U01DA050988, U01DA051039, U01DA041156, U01DA041025, U01DA041120, U01DA051038, U01DA041148, U01DA041093, U01DA041089, U24DA041123, U24DA041147. A full list of supporters is available at https://abcdstudy.org/federal-partners.html. A listing of participating sites and a complete listing of the study investigators can be found at https://abcdstudy.org/principal-investigators/. ABCD consortium investigators designed and implemented the study and/or provided data but did not necessarily participate in analysis or writing of this report. This manuscript reflects the views of the authors and may not reflect the opinions or views of the NIH or ABCD consortium investigators. The ABCD repository grows and changes over time. The ABCD data used in this report came from http://dx.doi.org/10.15154/1523041. DOIs can be found at nda.nih.gov.

## Notes

### Competing Interest Statement

The authors have declared no competing interest.

### Funding Statement

This study did not receive any funding

### Author Declarations

The study used ONLY openly available human data that were originally located at: http://dx.doi.org/10.15154/1523041.

