## Appendix for "The effect of multigeneration history of suicidality on offspring’s neurodevelopment outcomes: evidence from the Adolescent Brain and Cognitive Development (ABCD) cohort"

**Creation of composite demographic variable**

*Socio-economic Status (SES)* was derived from combined household income(Z1) and mean parental education(Z2). Then the factor analysis was used to obtain the factor loadings (β1, β2) and the eigenvalue of the primary factor(εf). SES score was calculated by the formula below^1^. It was then divided into high SES group and low SES group using 0 as the cut-off.

SES = (β1 * Z1 ＋ β2 * Z2) / εf

*Family history of suicide (FHoS)*

Q. Has ANY blood relative of your child ever attempted or death by suicide?

G1(Grandparents): maternal grandmother, maternal grandfather

paternal grandmother, paternal grandfather

G2(Parents): biological mother, biological father

**eMethods**

*Propensity Score Matched analysis (PSM)*

To account for the sample size differences, G1+G2-, G1-G2+ and G1+G2+ groups were combined as a new group, which was 1:1 matched to G1-G2- group on age, sex at birth, race/ethnicity, household marital status, parental highest education and household income. A one-to-one Nearest Neighborhood matching (NNM) algorithm was performed with a caliper of 0.2 and without replacement. Propensity scores were estimated using a logistic regression model.

*Generalised Estimated Equations (GEE)*

GEE represents an extension of the general linear model approach, allowing for correlations among site clusters. The 4-level family history of suicidality variable was analyzed as a categorical variable, with children without FHoS (G1-G2-) as the reference group. The association between child’s impulsivity and FHoS was assessed using GEE models with normal distribution and identity link function. The association between child’s SI and psychopathology and FHoS were assessed using similar GEE models with binomial distribution and logit link function.

**Supplement results**

| **GMV** | ***t*** | **P** | **P_FDR_** |
| --- | --- | --- | --- |
| superior segment of the circular sulcus of the insula | 12.891 | <0.001 | <0.001 |
| transverse frontopolar gyri and sulci | 10.405 | 0.001 | 0.025 |
| lateral occipito-temporal sulcus | 10.796 | 0.001 | 0.025 |
| posterior-dorsal part of the cingulate gyrus | 9.727 | 0.002 | 0.030 |
| planum polare of the superior temporal gyrus | 9.28 | 0.002 | 0.030 |
| precentral gyrus | 8.726 | 0.003 | 0.032 |
| vertical ramus of the anterior segment of the lateral sulcus | 8.739 | 0.003 | 0.032 |
| inferior occipital gyrus and sulcus | 8.476 | 0.004 | 0.034 |
| inferior temporal gyrus | 7.747 | 0.005 | 0.034 |
| posterior ramus of the lateral sulcus | 7.922 | 0.005 | 0.034 |
| temporal pole | 7.762 | 0.005 | 0.034 |
| orbital gyri | 7.119 | 0.008 | 0.049 |

1	Ren, ChunRong. Techniques for measuring the socio-economic status of students’ families. *Journal of Educational Studies* 2010; **5**: 77–82. [In Chinese]
